# Apraxic imitation deficits in Alzheimer’s disease are associated with altered dynamic connectivity

**DOI:** 10.1101/2024.04.05.24305320

**Authors:** Taylan D. Kuzu, Elena Jaeger, Anna K. Bonkhoff, Veronika Wunderle, Gérard N. Bischof, Kathrin Giehl, Maximilian H. T. Schmieschek, Özgür A. Onur, Frank Jessen, Gereon R. Fink, Alexander Drzezga, Peter H. Weiss

## Abstract

Apraxia is a common symptom in individuals with Alzheimer’s disease (AD). However, the neural mechanisms underlying apraxic deficits in AD remain elusive. Therefore, the current study focuses on the association between altered functional connectivity and apraxia in individuals with AD examining the hypothesis that apraxic deficits in AD result from dysfunction in praxis-related networks.

To this aim, we examined the association between changes in static and dynamic functional connectivity (FC) of resting-state networks and apraxia in AD. Resting-state functional MRI data was acquired in 13 patients with tau-and amyloid-positive AD, who underwent an extensive neuropsychological, motor and apraxia assessment, and 13 matched healthy control participants. The static and dynamic functional connectivity of resting state networks revealed by independent component analysis (ICA) were assessed and connectivity measures were correlated with apraxia scores.

Across all participants, we identified two distinct dynamic FC states using a sliding window approach. Patients with AD exhibited prolonged dwell times in the first state characterized by weaker connectivity and spent overall more time in this state, i.e., showed an increased fraction time in the first state. Apraxic deficits, especially deficits in imitating gestures, correlated significantly with the fraction time of both states as well as with the dwell time of the weakly connected dynamic state.

Data suggest that apraxic imitation deficits in AD are associated with dysfunction of praxis networks characterized by altered dynamic FC.

## Introduction

Apraxia is a disorder affecting cognitive motor functions related to gesture imitation, pantomiming object use and/or actual object use that cannot be (solely) attributed to basic motor deficits (Cubelli, 2017). Apraxic symptoms are observed in around 30-90% of individuals with Alzheimer’s disease (AD) (Lesourd et al., 2013; Stamenova et al., 2014). According to DSM-V, the diagnosis of AD, commonly associated with memory impairments and other cognitive deficits, requires the presence of additional impairments, e.g., apraxia (American Psychiatric Association, 2013). In AD, praxis deficits commonly manifest as limb apraxia, with the imitation of finger and hand gestures being especially impaired (Johnen et al., 2015). Despite the considerable limitations imposed by praxis disturbances on daily activities (Dovern et al., 2012), research on apraxia in individuals with AD has remained scarce. As a result, the neural mechanisms underlying apraxia in this neurodegenerative disease remain unclear.

To gain insights into the neural basis of apraxia, Watson and colleagues explored the static functional connectivity (FC) in stroke patients, employing resting-state functional magnetic resonance imaging (fMRI) (Watson et al., 2019). The authors found positive associations of pantomime and skilled tool use with interhemispheric connectivity involving cortical areas relevant for cognitive control, namely bilateral dorsolateral and anterior medial prefrontal cortices, and areas implicated in gesture production, namely the left posterior middle temporal gyrus. Static FC has also been assessed in individuals with AD (Kenny et al., 2012) to examine the associations of structural connectivity with FC and to compare FC patterns between different types of dementia, i.e., AD and Lewy body dementia. With the advent of the knowledge that cortical signals are not stationary even during rest, the analysis of the temporal characteristics of FC, i.e., dynamic FC, has gained importance (Allen et al., 2014). A dynamic FC study in AD patients demonstrated changes in connectivity strengths and temporal parameters (Jones et al., 2012). Specifically, Jones and colleagues reported group differences in dwell times of default mode network areas between AD patients and a healthy control group.

Here, we employed analyses of static and dynamic functional connectivity in patients with biomarker-verified AD pathology to disclose potential dysfunction of resting-state networks in AD patients with apraxia. Based on previous investigations of apraxia after stroke (Schmidt et al., 2022), we focused on three network domains. Specifically, the primary domain comprising the primary motor, visual and auditory networks; the premotor cortex, supplementary motor area (SMA) and cerebellar networks constituting the action domain and the intraparietal sulcus (IPS), inferior parietal lobe (IPL) as well as two fronto-parietal networks forming the parietal domain. For these resting-state networks, we computed parameters of static and dynamic connectivity and examined their association with the apraxic deficits observed in the current sample of patients.

We hypothesized that apraxia in patients with AD is related to dysfunctional connectivity patterns. Furthermore, we hypothesized that the apraxic imitation deficits typically observed in patients with AD are associated with changes in dynamic rather than static FC.

## Methods

### Participants

We recruited a total of 26 right-handed participants as determined by a score > 48 in the Edinburgh Handedness Inventory (Oldfield, 1971). Among those, there were 13 individuals diagnosed with dementia of AD type (11 males, 2 females; age: µ = 67.69 ± 12.59). The AD group was recruited from the interdisciplinary Center for Memory Disorders (ZfG; ‘Zentrum für Gedächtnisstörungen’) headed by the Departments of Neurology and Psychiatry of the University Hospital Cologne.

Patients were screened according to the following criteria: (i) age above 50 years; (ii) probable AD diagnosis according to National Institute on Aging and Alzheimer’s Association (NIA-AA) criteria (McKhann et al., 2011). The following inclusion criteria were applied: (i) dementia of AD type, as documented by clinical examination and positive biomarkers for Alzheimer pathology in cerebrospinal fluid (CSF) or positron emission tomography (PET) findings, i.e., positive Tau and Amyloid biomarkers (A+T+). (ii) apraxia operationalized by abnormal results in at least one of the applied apraxia tests.

The exclusion criteria were as follows: (i) suspected other forms of dementia than AD; other conditions responsible for cognitive or motor abnormalities, such as stroke, Parkinson’s disease, multiple sclerosis, epilepsy, or normal pressure hydrocephalus; incapacity to give informed consent; (iv) any contraindications to MRI.

As a healthy control group, 13 individuals (9 males, 4 females; age: µ = 68.00 ± 9.81) without neurological or psychiatric conditions were recruited. The study was approved by the local ethics committee of the Medical Faculty of the University of Cologne. All participants provided informed written consent. The study was conducted in accordance with the Declaration of Helsinki.

### Neuropsychological assessment

General cognitive functioning was assessed using the Mini-Mental State Examination (MMSE) (Folstein et al., 1975), the Dementia Detection test (DemTect) (Kalbe et al., 2004), and the Cologne Neuropsychological Screening for Stroke Patients (KöpSS) (Kaesberg et al., 2013). For the assessment of aphasia, the Aphasia Checklist in its abbreviated version (ACL-K) was used (Kalbe et al., 2002, 2005). Depressive symptoms were assessed using the Beck Depression Inventory II (BDI-II) (Beck et al., 1996) and the Montgomery-Åsberg Depression Rating Scale (MADRS) (Montgomery & Åsberg, 1979). Executive functioning was evaluated with the help of the Trail Making Tests A and B, with the ratio of B versus A controlling for basic motor deficits (Reitan, 1958).

### Motor assessment

Additionally, both groups underwent a comprehensive motor assessment, comprising the following tests: maximum grip strength using a vigorimeter (KLS Martin Group GmbH; Tuttlingen, Germany) (Desrosiers et al., 1995; Volz et al., 2016), maximum finger-tapping-frequency (Wang et al., 2009), the Purdue Pegboard Test (Reddon et al., 1988), the Jebsen-Taylor Hand Function Test (Jebsen et al., 1969) and the Action Research Arm Test (Lyle, 1981). Detailed descriptions of the tests can be found in the investigation on motor function across the life span by Wunderle and colleagues (2024).

### Apraxia assessment

The apraxia assessment was based on a range of previously established apraxia tests (Schmidt et al., 2022). It comprised the following tests: the Cologne Apraxia Screening (KAS) (Dovern et al., 2012; Weiss et al., 2013), the Goldenberg imitation tests for finger configurations and hand positions (Goldenberg, 1996), the Dementia Apraxia Test (DATE) (Johnen et al., 2016) and the De Renzi imitation test (De Renzi et al., 1980).

The KAS is a validated test for the assessment of apraxic symptoms in stroke patients and is also used effectively for the evaluation of apraxia in mild dementia (Johnen et al., 2018). It consists of four subtests that examine bucco-facial and arm/hand gestures in the context of two tasks, namely pantomime of object use and imitation.

### MRI acquisition

Magnetic resonance images were acquired using a Siemens MAGNETOM Prisma 3 Tesla scanner (Siemens Medical Solutions; Erlangen, Germany). For resting-state fMRI, participants were instructed to minimize motion throughout the 9.87-minute measurement period. They were asked not to fall asleep, keep their eyes open and fixate a white cross presented on the mirrored screen in front of them. Participants were allowed to let their thoughts wander. The following echo planar imaging (EPI) parameters were applied: repetition time = 0.8 s, echo time = 0.037 s, field of view = 208 mm, 72 axial slices, 2.0 mm^3^ isometric voxel size, flip angle = 52°, 740 volumes. Additionally, we recorded T1-images (MP RAGE, repetition time = 2.5 s, echo time = 2.22 ms, field of view = 241 mm, 208 axial slices, 0.94 mm^3^ isometric voxel size, flip angle = 7°) for EPI co-registration.

### Preprocessing of resting-state fMRI

Resting-state fMRI data were preprocessed using fMRIPrep version 23.1.4 (Esteban et al., 2019). Prior to preprocessing, the initial five volumes of each scan were discarded to ensure complete saturation of the magnetic field. For all remaining 735 volumes per subject, the following steps were applied: skull stripping, head motion estimation, spatiotemporal filtering, slice-timing correction, susceptibility distortion correction, co-registration, and normalization into MNI space. Importantly, the mean and maximum framewise displacement (FD) for each subject’s scan was computed as a measure of head motion (Jenkinson & Smith, 2001; Power et al., 2014). Further information about the preprocessing procedure can be found in the supplementary material. Finally, preprocessed fMRI images were smoothed with a Gaussian filter of 6 mm at full-width-at-half-maximum using Statistical Parametric Mapping (SPM 12; Wellcome Centre for Human Neuroimaging, London, UK), implemented in Matlab version 2023b (Mathworks Inc.; MA, USA). Before proceeding, we manually inspected outputs of head motion estimation and verified the absence of severe movement in all datasets, i.e., ensured that mean FD values were below 0.5 mm.

### Static and dynamic connectivity

For the analysis of functional connectivity (FC), we used the GIFT Toolbox Version 4.0 (https://trendscenter.org/software/gift/) implemented in SPM 12. Initially, a spatially constrained independent component analysis (ICA) was conducted to extract intrinsic components (Du & Fan, 2013; Lin et al., 2009), using the Neuromark_fMRI_2.1 template (Iraji et al., 2022), which was built upon datasets of over 57.000 individuals from various demographics (available in the GIFT Toolbox and at https://trendscenter.org/data/). Since the study primarily focused on (disturbed) connectivity in apraxia, our analysis focused on basic and complex motor systems. Based on previous research on the neuroanatomical and functional organization of the praxis networks and their pathophysiology (i.e., networks involved in apraxia; Martin et al., 2017; Schmidt & Weiss, 2021), as well as sensory and cognitive processing networks mentioned previously (Smith et al., 2009), we selected 10 intrinsic connectivity networks (ICNs), namely: (i) left primary sensorimotor cortex; (ii) right primary sensorimotor cortex; (iii) bilateral supplementary motor area (SMA); bilateral premotor cortex (PMC); (v) left cerebellum; (vi) right cerebellum; (vii) bilateral intraparietal sulcus (IPS); (viii) bilateral inferior parietal lobule (IPL); (ix) left fronto-parietal network; (x) right fronto-parietal network. As our motor and apraxia assessment are based on verbal commands and visual representations, we additionally included a visual and auditory ICN. We divided these networks into three network domains, namely the primary domain, the action domain, and the parietal domain (see Figure 1). Before further analysis, time courses were detrended, despiked using 3D despike, filtered by a low-pass filter with a high-frequency cut-off of 0.15 Hz, and normalized for variance (Rachakonda et al., 2007).

**Figure 1.**
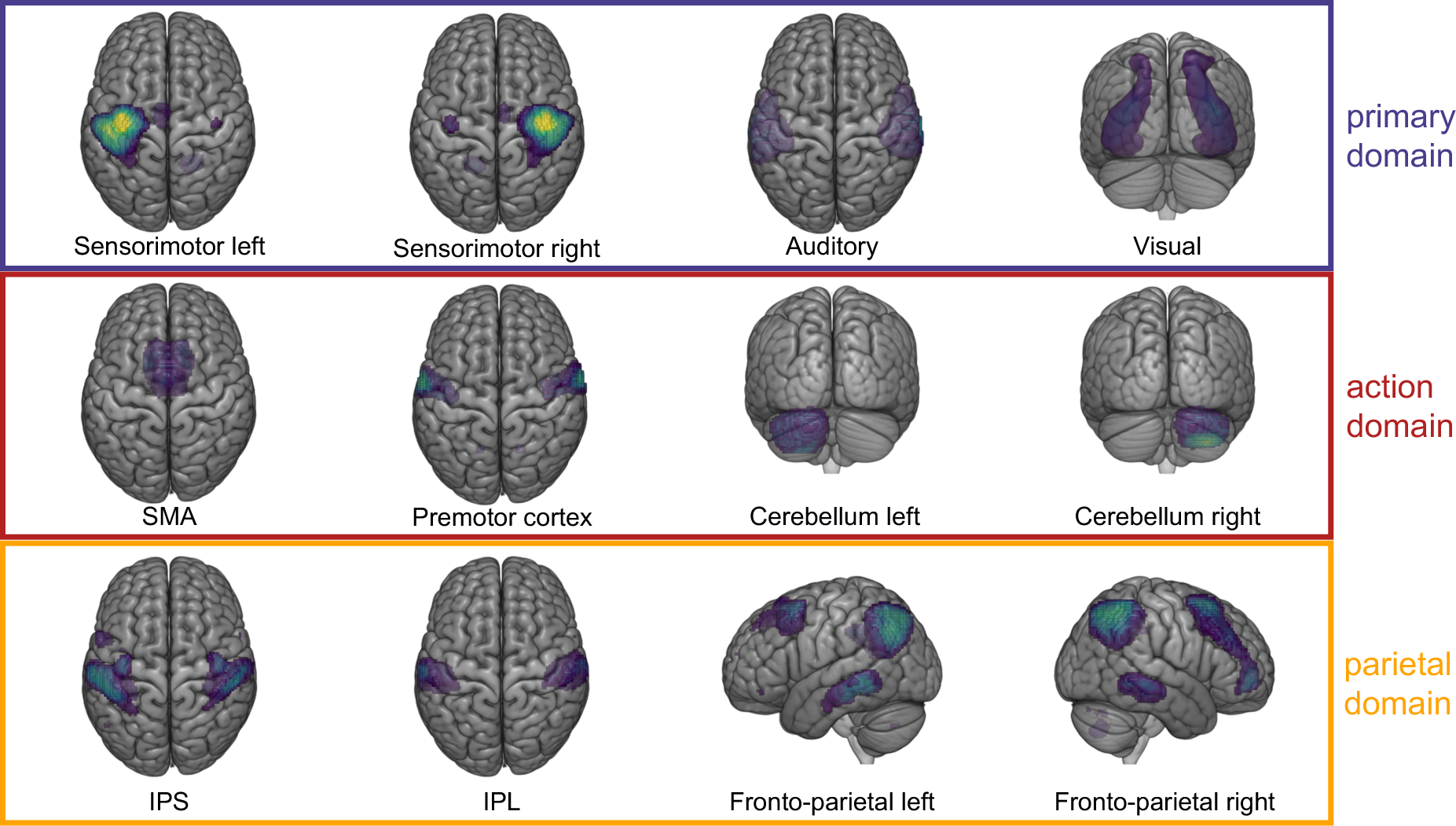
Intrinsic connectivity networks. The spatial maps of all 12 intrinsic connectivity networks (ICNs) for the entire study population (N = 26), consisting of 13 individuals diagnosed with Alzheimer’s disease (AD) and 13 healthy controls (HC). The ICNs were categorized into three different domains: sensorimotor domain (purple), complex motor domain (red), parietal domain (orange).

For the analysis of static FC, we employed the MANCOVAN toolbox as part of the GIFT Toolbox. We examined static FC within different networks and between networks. Age, sex, and FD were regressed out as covariates in all analysis. Connectivity was computed using mean Pearson’s pairwise correlations and reported as Fisher’s z-transformed correlations. Dynamic functional connectivity was analyzed using a sliding window approach (Allen et al., 2014; Calhoun et al., 2014; Damaraju et al., 2014; Sakoğlu et al., 2010) with a window size of 44 seconds, a Gaussian window alpha of three, and a window shift of 0.8 seconds (one repetition time). Thus, each measurement was divided into 657 individual windows. Within each window, dynamic FC was computed using the l1-regularized precision matrix. Age, sex, and FD were regressed out as covariates in all analyses. Similar to static FC, the correlations were Fisher z-transformed. To group similar connectivity patterns during the windows, we employed k-means clustering (Allen et al., 2014; Calhoun et al., 2014). Calculating an optimal cluster size, using (i) the silhouette score (Rousseeuw, 1987), and (ii) a state frequency of > 10%, revealed an optimal number of k = 2 states (see Figure S1).

### Statistics

For the statistical analysis of the data, we utilized Python version 3.11.3 and JASP version 0.18.3 (University of Amsterdam, Netherlands). For statistical analyses, a significance level of p < 0.05 was set. False-Discovery Rate (FDR)-correction at a significance level of p < 0.05 was applied to account for multiple comparisons (Benjamini & Hochberg, 1995). All results are depicted as mean ± standard deviation (SD). Concerning the neuropsychological and motor assessments, we examined group differences using independent Student’s t-tests.

### Static and dynamic connectivity analysis

Regarding static FC, we examined group differences within networks and connectivity pairs between networks using independent Student’s t-tests. For dynamic FC, we analyzed connectivity pairs between networks employing independent Student’s t-tests. We computed two analyses of variances (ANOVAs), each with the between-subject-factor group (levels: patients, controls) and the within-subject-factor state (levels: state 1, state 2) to evaluate the fraction time (i.e., the proportion of total time spent in one state) and the dwell time (i.e., the mean time spent in a given state without switching to another state). FDR-corrected post hoc t-tests were conducted in case of significant ANOVA results. Additionally, we investigated group differences in the number of transitions between states using an independent Student’s t-test.

### Correlating dynamic connectivity features with apraxia scores

We explored within the group of patients with AD whether differences in static and dynamic FC were associated with specific apraxic deficits as assessed by the KAS or with the severity of apraxia operationalized by the number of impaired apraxia tests. With regard to the KAS, we used the total score as well as the sum of subtests for each effector separately (bucco-facial and arm/hand) irrespective of the task, and the sum of subtests for both tasks (pantomime and imitation) regardless of the effectors for correlation analyses. We focused on measures from the preceding analysis of static and dynamic FC that revealed significant differences. Where normality was not met according to the Shapiro-Wilk test, we used Spearman correlations. Otherwise, we calculated Pearson correlations.

### Data availability

Data are available from the corresponding author upon reasonable request.

## Results

### Demographic and clinical characteristics

There were no significant differences between AD patients and controls regarding age (t (24) = –0.070, p = 0.945), sex (χ² (1, 26) = 0.217, p = 0.642), years of education (t (24) = 0.796, p = 0.434), laterality quotient of the EHI (t (24) = –1.686, p = 0.105), and framewise displacement (FD) (t (24) = –1.727, p = 0.097). An overview of the demographic information is illustrated in Table 1. All patients tested positive for amyloid-ß and tau with respect to at least one of the two diagnostic modalities, i.e., CSF or PET. An overview of the biomarker status for each patient can be found in the supplementary material (see Table S1). At the time of study conduction, 5 patients were taking acetyl cholinesterase inhibitors (donepezil = 4, galantamine = 1).

**Table 1.** Demographic characteristics in the study population.

| Category | Patients with AD<br>(N = 13) | Healthy controls<br>(N = 13) | p |
| --- | --- | --- | --- |
| Sex [m/ f] | 11/2 | 9/4 | 0.642 |
| Age [years] | 67.69 ± 12.59 | 68.00 ± 9.81 | 0.945 |
| Years of education [years] | 15.54 ± 2.79 | 16.39 ± 2.63 | 0.434 |
| Laterality quotient [%] | 96.08 ± 7.75 | 88.15 ± 15.07 | 0.105 |

The table depicts the demographic characteristics of the study population, divided into the groups of individuals with Alzheimer's disease (AD) and healthy controls. For the characteristics sex, age, years of education and laterality quotient according to the Edinburgh Handedness Inventory, descriptive data as well as statistical group differences are provided. For each group, descriptives are provided as means ± standard deviation (SD), and p-values of statistical group differences as calculated by Student's t-tests are reported for each of the subtests.

### Neuropsychological assessment

Within the group of 13 AD patients, 10 individuals exhibited symptoms of cognitive decline indicated by pathological results in the MMSE (i.e., ≤ 26/30 points). Among them, 9 patients displayed scores indicative of mild AD, while the remaining patient showed a score indicative of moderate AD. Further tests confirmed the symptoms of cognitive decline in AD patients (DemTect, KöpSS) and executive dysfunction (Trail Making Test). The groups did not differ significantly regarding depressive symptoms as evaluated by the BDI-II (t = 1.937, p_FDR_ = 0.072) and the MADRS (t = 1.883, p_FDR_ = 0.072). However, 3 patients scored above cut-off in the BDI-II and the MADRS indicating mild depressive symptoms. More detailed information about the neuropsychological assessment can be found in the supplementary material (see Table S2A).

### Motor assessment

Detailed results for the motor assessment are provided in the supplementary material (see Table S2B). After correction for multiple testing, no differences in motor function were observed between the two groups.

### Apraxia assessment

All 13 patients with AD exhibited apraxic deficits, as evidenced by pathological results in at least one apraxia test. In detail, 4 patients displayed deficits in 1 apraxia test, 3 patients showed deficits in 2 and 3 tests, respectively, and 1 patient exhibited pathological scores in 4 tests. The remaining 2 patients exhibited deficits in all 5 apraxia tests.

Since the KAS covers two tasks (imitation, pantomime) and two effectors (upper limb, bucco-facial), we focused on the KAS scores for the detailed analysis of the AD patients’ apraxic deficits. We evaluated the overall KAS score as well as the results of scores after categorization into tasks and effectors, i.e., praxis domains. In the current sample of AD patients, the mean overall KAS score was 73.54 ± 5.29 points. 11 of the 13 AD patients had pathological scores in the KAS. Regarding the praxis domains, the lowest mean score was observed for arm/ hand gestures (µ = 35.31 ± 4.46), with 11 patients showing abnormal results (see Kusch et al., 2018). Similar scores were achieved for the two tasks (pantomime: µ = 36.85 ± 3.36; imitation: µ = 36.77 ± 3.61).

7 and 9 AD patients were impaired in pantomime and imitation, respectively. The highest mean score for AD patients was observed for bucco-facial gestures (µ = 38.31 ± 2.25), with only 5 patients presenting abnormal values. Detailed information on the results of the apraxia assessment can be found in Table 2.

**Table 2.** Results of the apraxia assessment in the current sample of AD patients (N = 13).

| Subtest | Number of impaired AD patients | Mean and SD for the AD patient group |
| --- | --- | --- |
| KAS total | 9 | 73.54 ± 5.29 |
| KAS pantomime | 7 | 36.85 ± 3.36 |
| KAS imitation | 9 | 36.77 ± 3.61 |
| KAS bucco-facial | 5 | 38.31 ± 2.25 |
| KAS arm/hand | 11 | 35.31 ± 4.46 |
| Goldenberg imitation finger | 6 | 15.92 ± 3.25 |
| Goldenberg imitation hand | 6 | 17.23 ± 2.46 |
| Dementia Apraxia Test (DATE) | 6 | 45.46 ± 9.59 |
| De Renzi imitation test | 3 | 58.39 ± 8.93 |

The table illustrates the results for each subtest comprising the apraxia assessment as well as the effector and task domains of the KAS. Cut-off values for each domain of the KAS (i.e., $\leq 38/40$ points) have been reported in (Kusch et al., 2018). Descriptive data are provided as means ± standard deviation (SD) as well as the number of affected patients per test.

### Static and dynamic functional connectivity

There were no statistically significant group differences within the spatial maps of any network component after FDR-correction. Thus, the static connectivity within the ICNs did not significantly differ between AD patients and controls.

Across all participants, analysis of between-network static FC revealed positive correlations between sensorimotor, SMA, PMC, IPS and IPL network components. In contrast, the correlation of the auditory, visual, and right fronto-parietal network components with the other network components were weak. The cerebellar components and the left fronto-parietal network component were negatively connected with the other components. A depiction of the static connectivity matrix can be found in Figure 2.

**Figure 2.**
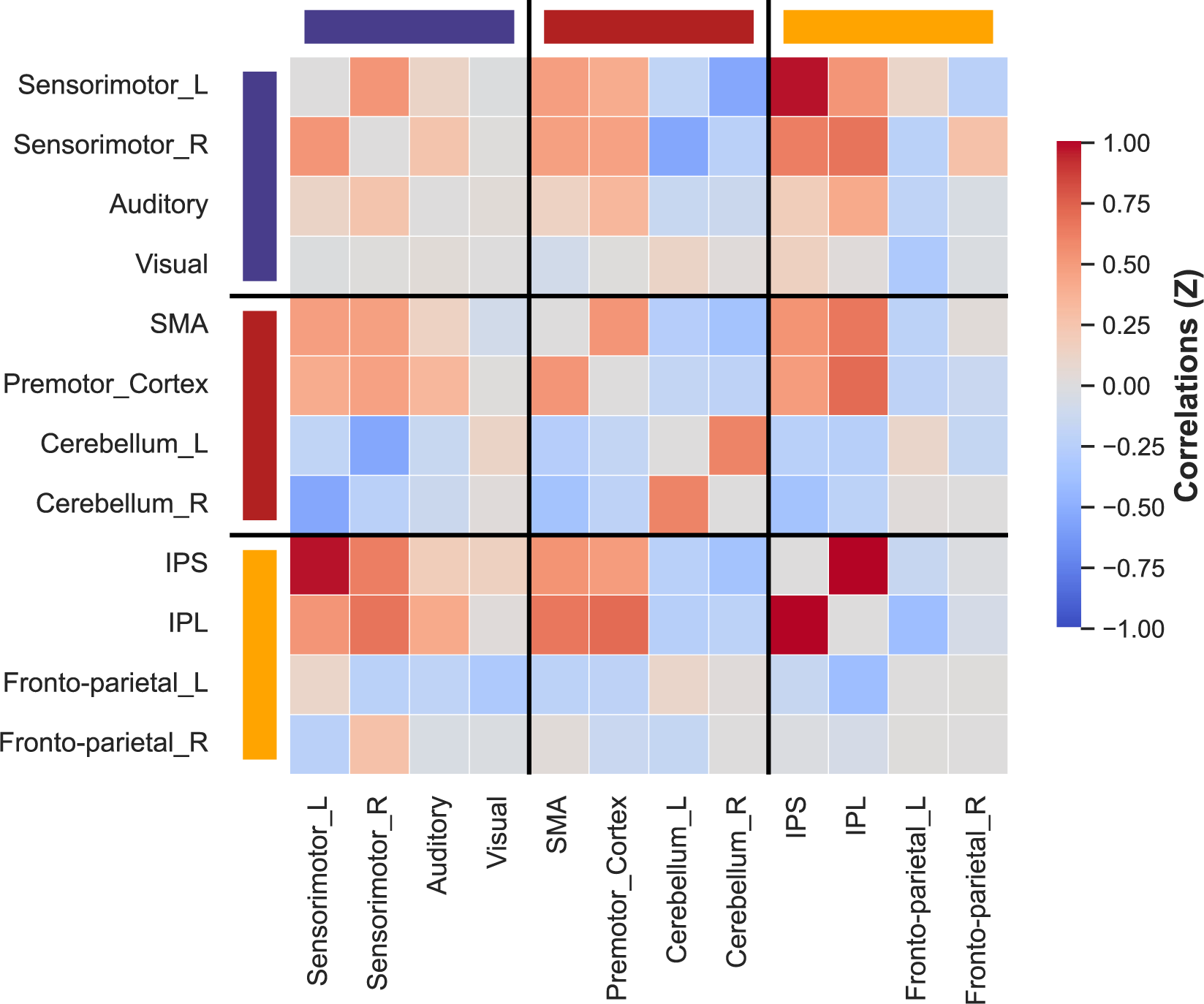
Static connectivity matrix. The static connectivity matrix averaged over all patients with Alzheimer’s disease (AD) and healthy controls (HC), depicting pairs of high (red) and low (blue) connectivity between all intrinsic connectivity networks (ICNs). The colored bars at the top and left side correspond to the three domains presented in Figure 1 (purple: sensorimotor domain, red: complex motor domain, orange: parietal domain). There were no significant group differences regarding static connectivity between ICNs.

The dynamic FC analysis revealed two states characterized by similar patterns regarding strength of connectivity between individual network components. In the first state, these positive and negative associations were markedly weakened, while in the second state, they were more pronounced. Correlations between connectivity values of the two dynamic states and the static connectivity revealed that the connectivity of the second dynamic state more closely reflected static connectivity compared to the first dynamic state. There were no statistical group differences regarding the dynamic FC of both states after FDR-correction. A depiction of the dynamic connectivity matrices per group can be found in Figure 3.

**Figure 3.**
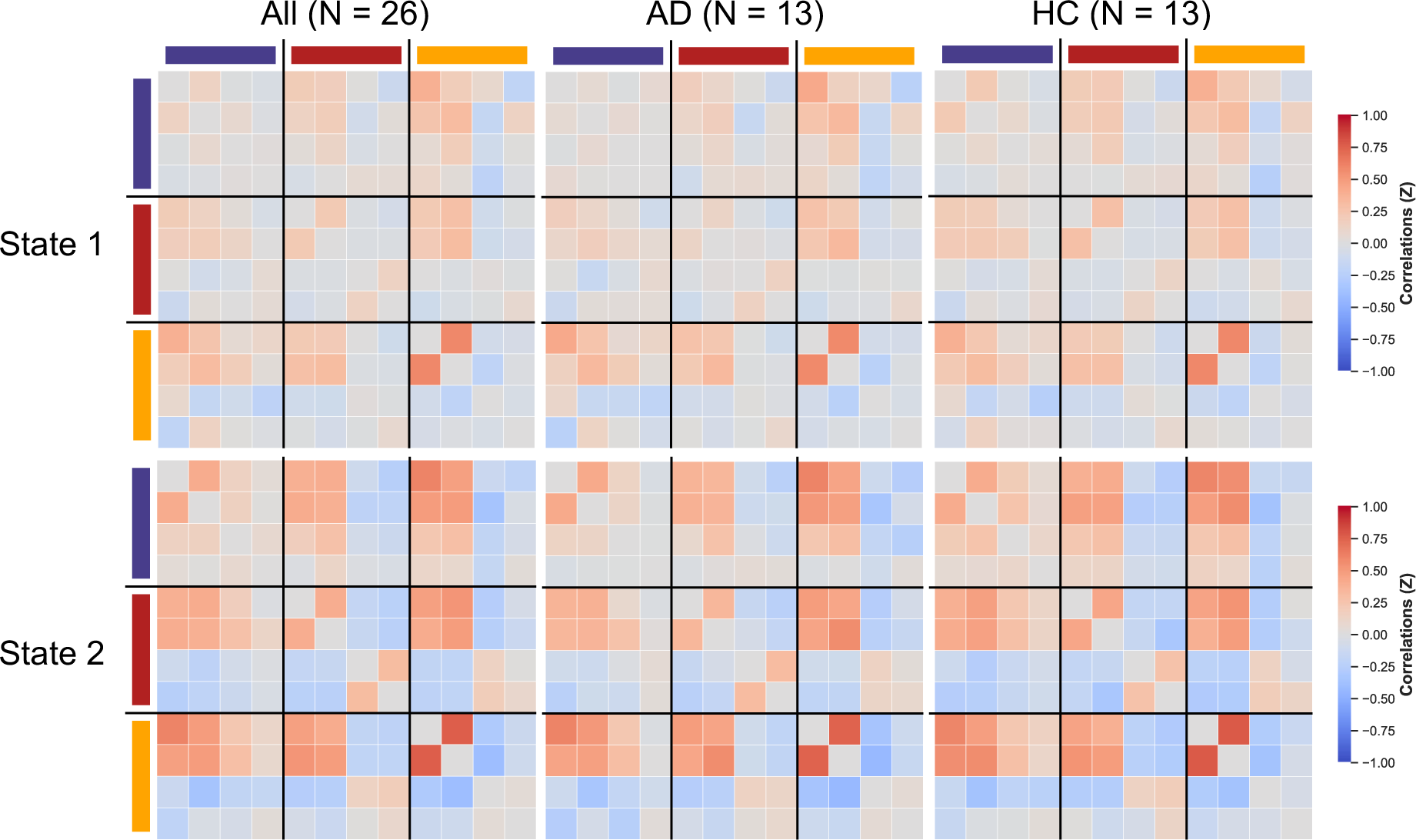
Dynamic connectivity matrices. The dynamic connectivity matrices of the first and second dynamic state over the entire study population, as well as for patients with Alzheimer’s disease (AD) and healthy controls (HC) separately. Depicted are pairs of high (red) and low (blue) connectivity between all intrinsic connectivity networks (ICNs). The colored bars at the top and left side correspond to the three domains presented in Figure 1. State 1 resembled the static connectivity matrix the most. There were no significant group differences regarding dynamic connectivity in the first or second state between ICNs.

Considering temporal characteristics, the first state with weaker connectivity occurred 10,420 times across the entire measurement of all participants (61.00%), while the second state with stronger associations occurred 6,662 times (39.00%). Figure 4A illustrates that AD patients transitioned less frequently between states compared to controls. However, the difference was not statistically significant (t (24) = –1.590, p = 0.125). Figure 4B presents the mean fraction times per state and group. The statistical analysis using an ANOVA revealed a significant effect of the variable states (F (1, 50) = 7.439, p < 0.01, η^2^ = 0.125) and a significant state x group interaction (F (1, 50) = 4.213, p < 0.05, η^2^ = 0.071) for fraction time. Post-hoc t-tests indicated that AD patients had a significantly higher fraction time in the first compared to the second state (t (25) = 3.380, p_FDR_ < 0.01), i.e., AD patients showed a higher proportion of total time spent in the first state. In contrast, no such difference for the fraction time between the two states was observed for controls (t (25) = 0.477, p = 1.000). For dwell times, i.e., the mean time spent in a given state without switching to another state (see Figure 4C), an ANOVA revealed a significant effect of the variable states (F (1, 48) = 6.355, p < 0.05, η^2^ = 0.108) and the interaction of the variables states and groups (F (1, 48) = 3.507, p < 0.05, η^2^ = 0.069). Post-hoc t-tests indicated a significantly longer dwell time in the first compared to the second state for AD patients (t (25) = 3.041, p_FDR_ < 0.05), but not for controls (t (25) = 0.469, p_FDR_ = 1.000).

**Figure 4.**
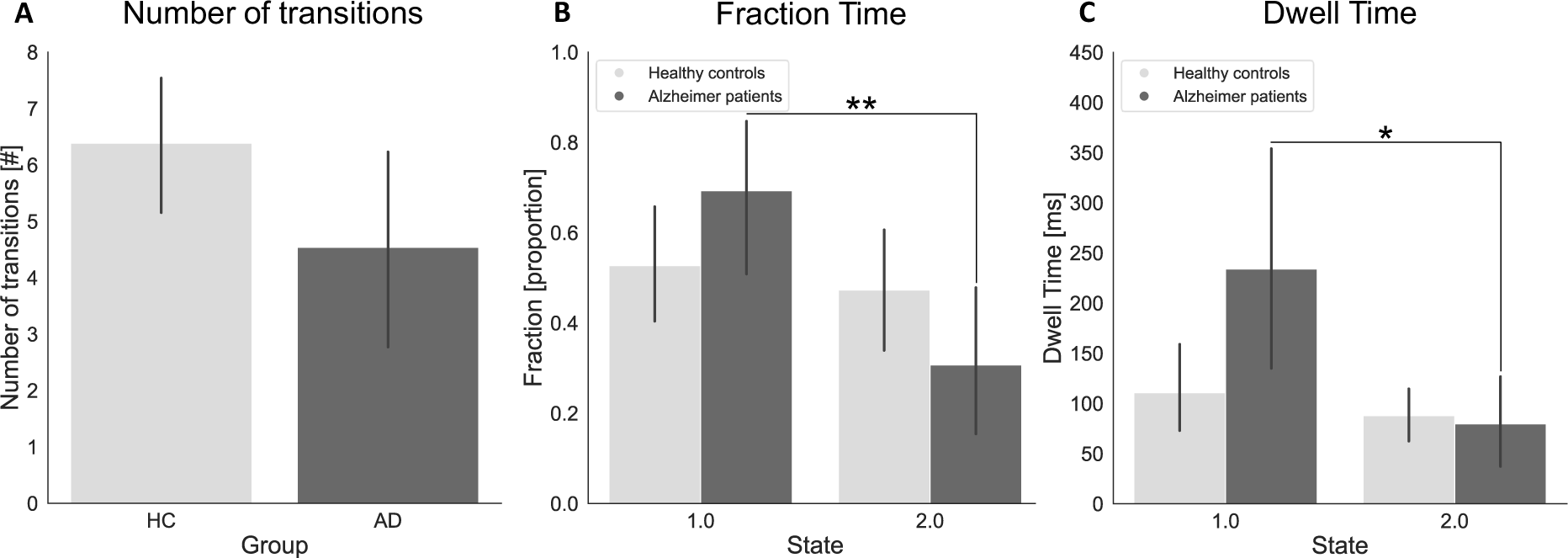
Dynamic functional connectivity analysis results. The results of the dynamic functional connectivity (FC) analysis regarding fraction and dwell time as well as the number of transitions between for the group of patients with Alzheimer’s disease (AD) and healthy controls (HC). **A.** Number of transitions. Depicted are the number of transitions for the HC (light gray) and AD (dark gray) group. **B.** Fraction times. Depicted are the fraction times for the HC (light gray) and AD (dark gray) group and separately per dynamic state. **C.** Dwell times. Depicted are the dwell times for the HC (light gray) and AD (dark gray) group and separately per dynamic state. Asterisks indicate statistically significant differences after post-hoc t-tests: * = p < 0.05, ** = p < 0.01

### Associations between dynamic connectivity features and apraxia scores

No significant associations were found between the static FC measures and apraxia scores after FDR-correction.

We correlated temporal dynamic FC measures (i.e., fraction and dwell time) of the two dynamic states with the apraxia scores and controlled for the temporal difference (in days) between the apraxia assessment and the MRI measurement as well as for cognitive decline (operationalized by the MMSE). The KAS imitation domain scores were negatively associated with the mean fraction times of the first state and positively associated with the mean fraction times of the second state (see Table 3A). While there were no significant associations between the dwell times of the second state and apraxia scores after correction for multiple comparisons, poorer KAS imitation domain scores were associated with a longer dwell time spent in the first dynamic state (see Table 3B).

**Table 3A.** Correlation analysis between fraction times and apraxia scores. Correlation Fraction Time State 1 Fraction Time State 2.

| Correlation | Fraction Time State 1 |  | Fraction Time State 2 |  |
| --- | --- | --- | --- | --- |
|  | Coefficient | P<sub>FDR</sub> | Coefficient | p<sub>FDR</sub> |
| KAS sum | −0.182 | 0.710 | 0.182 | 0.710 |
| KAS pantomime | 0.304 | 0.710 | −0.304 | 0.710 |
| **KAS imitation** | **−0.828** | **< 0.05\*** | **0.828** | **< 0.05\*** |
| KAS bucco-facial | 0.085 | 0.804 | −0.085 | 0.804 |
| KAS arm/hand | −0.213 | 0.710 | 0.213 | 0.710 |

**Table 3B.** Correlation analysis between dwell times and apraxia scores. Correlation Dwell Time State 1 Dwell Time State 2.

| Correlation | Dwell Time State 1 |  | Dwell Time State 2 |  |
| --- | --- | --- | --- | --- |
|  | Coefficient | P<sub>FDR</sub> | Coefficient | p<sub>FDR</sub> |
| KAS sum | −0.289 | 0.584 | −0.109 | 0.901 |
| KAS pantomime | 0.080 | 0.815 | −0.510 | 0.480 |
| **KAS imitation** | **−0.743** | **< 0.05\*** | 0.669 | 0.294 |
| KAS bucco-facial | −0.044 | 0.898 | −0.160 | 0.901 |
| KAS arm/hand | −0.361 | 0.584 | −0.133 | 0.901 |

## Discussion

In the present study, we investigated - for the first time - the static and dynamic FC during rest in patients with AD and apraxia. In apraxic AD patients, measures of dynamic connectivity, but not static connectivity parameters were associated with imitation deficits, suggesting that apraxic imitation deficits in AD are associated with dysfunction of praxis networks characterized by altered dynamic FC.

### Altered dynamic connectivity in Alzheimer’s disease

The analysis of dynamic FC in the current sample of AD patients and healthy controls suggested a division into two states, namely a weaker connected - more integrated - and a stronger connected - more segregated - configuration. Importantly, group differences were revealed with respect to the temporal characteristics of dynamic FC. AD patients spent significantly more time in the more integrated state than in the more segregated state and remained longer in that state. Moreover, apraxic AD patients switched – albeit not significantly - less often between the two states during the resting-state fMRI session. In contrast, controls spent similar durations in both states. Our results are consistent with previous studies that investigated dynamic FC in AD and found group differences in the temporal patterns of dynamic connectivity parameters (Jones et al., 2012). Thus, altered dynamic connectivity seems to be a hallmark of AD. Notably, our study is the first revealing an association between altered dynamic connectivity and apraxic imitation deficits in AD. Furthermore, the observed patterns of segregation and integration align with reports from the literature indicating that patients with neurodegenerative diseases spend more time in states featuring more integration (Kim et al., 2017; Hensel et al., 2023).

There is only one previous study on FC in apraxia. In this work, Watson and colleagues (2019) analyzed static connectivity in chronic left-hemisphere stroke patients using a seed-based approach. They found positive associations between performance in a pantomime task and static interhemispheric connectivity of the left superior parietal cortex, left postcentral gyrus, and left posterior medial temporal gyrus with their corresponding homologous areas in the right hemisphere. Notably, our study did not reveal any group differences for static FC, but for the temporal parameters of dynamic FC. Thus, it seems that differential alterations in static and dynamic FC may underly apraxic deficits in stroke patients and patients with AD, respectively. These differential associations of apraxic deficits with static or dynamic connectivity changes could be related to the different time courses of the two diseases (stroke: sudden onset; AD: slowly progressing) or their different etiology (stroke: ischemia or hemorrhage; AD: neurodegenerative process).

### Imitation deficits in Alzheimer’s disease

Within the group of AD patients, the fraction times of the two states and the dwell times of the low connectivity state 1 correlated with apraxic imitation deficits as assessed by the KAS. More severe impairments in imitation (i.e., lower KAS imitation scores) were associated with more time spent (i.e., longer fraction and dwell times) in the weaker connected, more integrated state, while less severe impairments in gesture imitation (i.e., higher KAS imitation scores) were associated with less time spent (i.e., shorter fraction times) in the stronger connected, more segregated state. Thus, our results demonstrated a close association between the temporal measures of dynamic FC and the clinically observed apraxic imitation deficits in AD. This aligns with previous research indicating that apraxic deficits in patients with AD are particularly evident for the imitation of limb gestures (Johnen et al., 2016). To the best of our knowledge, this is the first study to investigate dynamic connectivity in patients with AD and apraxia, demonstrating associations between transient connectivity alterations and apraxic symptoms.

### Limitations

This study also suffers from limitations. The current study population was relatively small, i.e., 13 AD patients with apraxia and 13 matched healthy participants. Notably, the current patient population consisted only of individuals, who were diagnosed with AD based on positive biomarker status (i.e., A+ T+ in either CSF or PET). Despite the small sample size, we were able to demonstrate robust and statistically significant group differences in dynamic FC. Our findings, along with prior investigations involving similar patient sample sizes (Greicius et al., 2004), suggest that even a relatively low number of patients can be sufficient for the analysis of transient changes in FC using resting-state fMRI measurements.

The patient sample in this study predominantly exhibited mild cognitive deficits. For future research, it would be interesting to take a closer look at patient populations in the individual distinct stages of AD, investigating whether there are differences in the association of connectivity with apraxia scores for the respective subpopulations.

### Summary

In summary, the present study replicates previous characteristic findings in the analysis of dynamic FC in neurodegenerative diseases and provides new insights into the crucial role of dynamic connectivity patterns regarding apraxic imitation deficits in AD, in addition to the conventional analysis of static FC. Associations between dynamic connectivity parameters and apraxia measures were identified. In contrast, we found no association with any parameter of static connectivity. This suggests that apraxic imitation deficits in AD result from dysfunction of praxis networks characterized by altered dynamic FC.

## References

1. Allen, E. A., Damaraju, E., Plis, S. M., Erhardt, E. B., Eichele, T., & Calhoun, V. D. (2014). Tracking Whole-Brain Connectivity Dynamics in the Resting State. Cerebral Cortex, 24(3), 663–676. 10.1093/cercor/bhs352

2. American Psychiatric Association. (2013). Diagnostic and Statistical Manual of Mental Disorders (Fifth Edition). American Psychiatric Association. 10.1176/appi.books.9780890425596

3. Beck, A. T., Steer, R. A., & Brown, G. (1996). Beck Depression Inventory–II [dataset]. 10.1037/t00742-000

4. Benjamini, Y., & Hochberg, Y. (1995). Controlling the False Discovery Rate: A Practical and Powerful Approach to Multiple Testing. Journal of the Royal Statistical Society: Series B (Methodological), 57(1), 289–300. 10.1111/j.2517-6161.1995.tb02031.x

5. Calhoun, V. D., Miller, R., Pearlson, G., & Adalı, T. (2014). The Chronnectome: Time-Varying Connectivity Networks as the Next Frontier in fMRI Data Discovery. Neuron, 84(2), 262–274. 10.1016/j.neuron.2014.10.015

6. Cubelli, R. (2017). Definition: Apraxia. Cortex, 93, 227. 10.1016/j.cortex.2017.03.012

7. Damaraju, E., Allen, E. A., Belger, A., Ford, J. M., McEwen, S., Mathalon, D. H., Mueller, B. A., Pearlson, G. D., Potkin, S. G., Preda, A., Turner, J. A., Vaidya, J. G., van Erp, T. G., & Calhoun, V. D. (2014). Dynamic functional connectivity analysis reveals transient states of dysconnectivity in schizophrenia. NeuroImage. Clinical, 5, 298–308. 10.1016/j.nicl.2014.07.003

8. De Renzi, E. (1989). Apraxia. In F. Boller & J. Grafman, Handbook of neuropsychology (pp. 245–263). Elsevier Science Publishers.

9. De Renzi, E., Motti, F., & Nichelli, P. (1980). Imitating gestures. A quantitative approach to ideomotor apraxia. Archives of Neurology, 37(1), 6–10. 10.1001/archneur.1980.00500500036003

10. Desrosiers, J., Hébert, R., Bravo, G., & Dutil, E. (1995). Comparison of the Jamar dynamometer and the Martin vigorimeter for grip strength measurements in a healthy elderly population. Journal of Rehabilitation Medicine, 27(3), 137–143. 10.2340/165019779527137143

11. Dovern, A., Fink, G. R., & Weiss, P. H. (2012). Diagnosis and treatment of upper limb apraxia. Journal of Neurology, 259(7), 1269–1283. 10.1007/s00415-011-6336-y

12. Du, Y., & Fan, Y. (2013). Group information guided ICA for fMRI data analysis. NeuroImage, 69, 157–197. 10.1016/j.neuroimage.2012.11.008

13. Esteban, O., Markiewicz, C. J., Blair, R. W., Moodie, C. A., Isik, A. I., Erramuzpe, A., Kent, J. D., Goncalves, M., DuPre, E., Snyder, M., Oya, H., Ghosh, S. S., Wright, J., Durnez, J., Poldrack, R. A., & Gorgolewski, K. J. (2019). fMRIPrep: A robust preprocessing pipeline for functional MRI. Nature Methods, 16(1), Article 1. 10.1038/s41592-018-0235-4

14. Folstein, M. F., Folstein, S. E., & McHugh, P. R. (1975). “Mini-mental state”: A practical method for grading the cognitive state of patients for the clinician. Journal of Psychiatric Research, 12(3), 189–198. 10.1016/0022-3956(75)90026-6

15. Goldenberg, G. (1996). Defective imitation of gestures in patients with damage in the left or right hemispheres. Journal of Neurology, Neurosurgery, and Psychiatry, 61(2), 176–180. 10.1136/jnnp.61.2.176

16. He, Y., Wang, L., Zang, Y., Tian, L., Zhang, X., Li, K., & Jiang, T. (2007). Regional coherence changes in the early stages of Alzheimer’s disease: A combined structural and resting-state functional MRI study. NeuroImage, 35(2), 488–500. 10.1016/j.neuroimage.2006.11.042

17. Iraji, A., Fu, Z., Faghiri, A., Duda, M., Chen, J., Rachakonda, S., DeRamus, T., Kochunov, P., Adhikari, B. M., Belger, A., Ford, J. M., Mathalon, D. H., Pearlson, G. D., Potkin, S. G., Preda, A., Turner, J. A., Erp, T. G. M. van, Bustillo, J. R., Yang, K.,…Calhoun, V. D. (2022). Canonical and Replicable Multi-Scale Intrinsic Connectivity Networks in 100k+ Resting-State fMRI Datasets (p. 2022.09.03.506487). bioRxiv. 10.1101/2022.09.03.506487

18. Jebsen, R. H., Taylor, N., Trieschmann, R. B., Trotter, M. J., & Howard, L. A. (1969). An objective and standardized test of hand function. Archives of Physical Medicine and Rehabilitation, 50(6), 311–319.

19. Jenkinson, M., & Smith, S. (2001). A global optimisation method for robust affine registration of brain images. Medical Image Analysis, 5(2), 143–156. 10.1016/S1361-8415(01)00036-6

20. Johnen, A., Frommeyer, J., Modes, F., Wiendl, H., Duning, T., & Lohmann, H. (2016). Dementia Apraxia Test (DATE): A Brief Tool to Differentiate Behavioral Variant Frontotemporal Dementia from Alzheimer’s Dementia Based on Apraxia Profiles. Journal of Alzheimer’s Disease: JAD, 49(3), 593–605. 10.3233/JAD-150447

21. Johnen, A., Reul, S., Wiendl, H., Meuth, S. G., & Duning, T. (2018). Apraxia profiles—A single cognitive marker to discriminate all variants of frontotemporal lobar degeneration and Alzheimer’s disease. *Alzheimer’s & Dementia: Diagnosis*, Assessment & Disease Monitoring, 10(1), 363–371. 10.1016/j.dadm.2018.04.002

22. Johnen, A., Tokaj, A., Kirschner, A., Wiendl, H., Lueg, G., Duning, T., & Lohmann, H. (2015). Apraxia profile differentiates behavioural variant frontotemporal from Alzheimer’s dementia in mild disease stages. Journal of Neurology, Neurosurgery & Psychiatry, 86(7), 809–815. 10.1136/jnnp-2014-308773

23. Jones, D. T., Vemuri, P., Murphy, M. C., Gunter, J. L., Senjem, M. L., Machulda, M. M., Przybelski, S. A., Gregg, B. E., Kantarci, K., Knopman, D. S., Boeve, B. F., Petersen, R. C., & Jack, C. R. (2012). Non-Stationarity in the “Resting Brain’s” Modular Architecture. PLoS ONE, 7(6), e39731. 10.1371/journal.pone.0039731

24. Kaesberg, S., Kalbe, E., Finis, J., Kessler, J., & Fink, G. R. (2013). KöpSS: Kölner Neuropsychologisches Screening für Schlaganfall-Patienten. Hogrefe.

25. Kalbe, E., Kessler, J., Calabrese, P., Smith, R., Passmore, A. P., Brand, M., & Bullock, R. (2004). DemTect: A new, sensitive cognitive screening test to support the diagnosis of mild cognitive impairment and early dementia. International Journal of Geriatric Psychiatry, 19(2), 136–143. 10.1002/gps.1042

26. Kalbe, E., Reinhold, N., Brand, M., & Kessler, J. (2002). The short aphasia-check-list: An economical screening for detecting aphasia. Eur. J. Neurol, 9, 209–210.

27. Kalbe, E., Reinhold, N., Brand, M., Markowitsch, H. J., & Kessler, J. (2005). A new test battery to assess aphasic disturbances and associated cognitive dysfunctions—German normative data on the aphasia check list. Journal of Clinical and Experimental Neuropsychology, 27(7), 779–794. 10.1080/13803390490918273

28. Kenny, E. R., Blamire, A. M., Firbank, M. J., & O’Brien, J. T. (2012). Functional connectivity in cortical regions in dementia with Lewy bodies and Alzheimer’s disease. Brain, 135(2), 569–581. 10.1093/brain/awr327

29. Kusch, M., Schmidt, C. C., Göden, L., Tscherpel, C., Stahl, J., Saliger, J., Karbe, H., Fink, G. R., & Weiss, P. H. (2018). Recovery from apraxic deficits and its neural correlate. Restorative Neurology and Neuroscience, 36(6), 669–678. 10.3233/RNN-180815

30. Lesourd, M., Le Gall, D., Baumard, J., Croisile, B., Jarry, C., & Osiurak, F. (2013). Apraxia and Alzheimer’s disease: Review and perspectives. Neuropsychology Review, 23(3), 234–256. 10.1007/s11065-013-9235-4

31. Lin, Q., Liu, J., Zheng, Y., Liang, H., & Calhoun, V. D. (2009). Semiblind spatial ICA of fMRI using spatial constraints. Human Brain Mapping, 31(7), 1076–1088. 10.1002/hbm.20919

32. Lyle, R. C. (1981). A performance test for assessment of upper limb function in physical rehabilitation treatment and research. International Journal of Rehabilitation Research. Internationale Zeitschrift Fur Rehabilitationsforschung. Revue Internationale De Recherches De Readaptation, 4(4), 483–492. 10.1097/00004356-198112000-00001

33. Martin, M., Hermsdörfer, J., Bohlhalter, S., & Weiss, P. H. (2017). [Networks involved in motor cognition: Physiology and pathophysiology of apraxia]. Der Nervenarzt, 88(8), 858– 865. 10.1007/s00115-017-0370-7

34. McKhann, G. M., Knopman, D. S., Chertkow, H., Hyman, B. T., Jack, C. R., Kawas, C. H., Klunk, W. E., Koroshetz, W. J., Manly, J. J., Mayeux, R., Mohs, R. C., Morris, J. C., Rossor, M. N., Scheltens, P., Carrillo, M. C., Thies, B., Weintraub, S., & Phelps, C. H. (2011). The diagnosis of dementia due to Alzheimer’s disease: Recommendations from the National Institute on Aging-Alzheimer’s Association workgroups on diagnostic guidelines for Alzheimer’s disease. Alzheimer’s & Dementia, 7(3), 263–269. 10.1016/j.jalz.2011.03.005

35. Montgomery, S. A., & Åsberg, M. (1979). A New Depression Scale Designed to be Sensitive to Change. British Journal of Psychiatry, 134(4), 382–389. 10.1192/bjp.134.4.382

36. Oldfield, R. C. (1971). The assessment and analysis of handedness: The Edinburgh inventory. Neuropsychologia, 9(1), 97–113. 10.1016/0028-3932(71)90067-4

37. Osiurak, F., & Rossetti, Y. (2017). Definition: Limb apraxia. Cortex, 93, 228. 10.1016/j.cortex.2017.03.010

38. Power, J. D., Mitra, A., Laumann, T. O., Snyder, A. Z., Schlaggar, B. L., & Petersen, S. E. (2014). Methods to detect, characterize, and remove motion artifact in resting state fMRI. NeuroImage, 84, 320–341. 10.1016/j.neuroimage.2013.08.048

39. Rachakonda, S., Egolf, E., Correa, N., & Calhoun, V. (2007). Group ICA of fMRI toolbox (GIFT) manual.

40. Reddon, J. R., Gill, D. M., Gauk, S. E., & Maerz, M. D. (1988). Purdue Pegboard: Test-Retest Estimates. Perceptual and Motor Skills, 66(2), 503–506. 10.2466/pms.1988.66.2.503

41. Reitan, R. M. (1958). Validity of the Trail Making Test as an Indicator of Organic Brain Damage. Perceptual and Motor Skills, 8(3), 271–276. 10.2466/pms.1958.8.3.271

42. Rousseeuw, P. J. (1987). Silhouettes: A graphical aid to the interpretation and validation of cluster analysis. Journal of Computational and Applied Mathematics, 20, 53–65. 10.1016/0377-0427(87)90125-7

43. Sakoğlu, U., Pearlson, G. D., Kiehl, K. A., Wang, Y. M., Michael, A. M., & Calhoun, V. D. (2010). A method for evaluating dynamic functional network connectivity and task-modulation: Application to schizophrenia. Magma (New York, N.Y.), 23(5–6), 351–366. 10.1007/s10334-010-0197-8

44. Schmidt, C. C., Achilles, E. I. S., Fink, G. R., & Weiss, P. H. (2022). Distinct cognitive components and their neural substrates underlying praxis and language deficits following left hemisphere stroke. Cortex, 146, 200–215. 10.1016/j.cortex.2021.11.004

45. Schmidt, C., & Weiss, P. (2021). The Cognitive Neuroscience of Apraxia. In Encyclopedia of Behavioral Neuroscience, 2nd edition (Second Edition) (pp. 668–677). 10.1016/B978-0-12-819641-0.00143-2

46. Smith, S. M., Fox, P. T., Miller, K. L., Glahn, D. C., Fox, P. M., Mackay, C. E., Filippini, N., Watkins, K. E., Toro, R., Laird, A. R., & Beckmann, C. F. (2009). Correspondence of the brain’s functional architecture during activation and rest. Proceedings of the National Academy of Sciences, 106(31), 13040–13045. 10.1073/pnas.0905267106

47. Stamenova, V., Roy, E. A., & Black, S. E. (2014). A model-based approach to limb apraxia in Alzheimer’s disease. Journal of Neuropsychology, 8(2), 246–268. 10.1111/jnp.12023

48. Volz, L. J., Rehme, A. K., Michely, J., Nettekoven, C., Eickhoff, S. B., Fink, G. R., & Grefkes, C. (2016). Shaping Early Reorganization of Neural Networks Promotes Motor Function after Stroke. Cerebral Cortex, 26(6), 2882–2894. 10.1093/cercor/bhw034

49. Wang, L. E., Fink, G. R., Dafotakis, M., & Grefkes, C. (2009). Noradrenergic stimulation and motor performance: Differential effects of reboxetine on movement kinematics and visuomotor abilities in healthy human subjects. Neuropsychologia, 47(5), 1302–1312. 10.1016/j.neuropsychologia.2009.01.024

50. Watson, C. E., Gotts, S. J., Martin, A., & Buxbaum, L. J. (2019). Bilateral functional connectivity at rest predicts apraxic symptoms after left hemisphere stroke. NeuroImage. Clinical, 21, 101526. 10.1016/j.nicl.2018.08.033

51. Weiss, P. H., Kalbe, E., Kessler, J., Fink, G. R., Binder, E., Hesse, M. D., & Scherer, A. (2013). Kölner apraxie screening. Hogrefe.

52. Wunderle V., Kuzu T. D., Tscherpel C., Fink G. R., Grefkes C., Weiss P. H. (2024). Age-and gender-related changes in motor functions: A Comprehensive Assessment and Component Analysis. medRxiv; ID#: MEDRXIV/2024/305334

