## Supplementary Material for "Apraxic imitation deficits in Alzheimer’s disease are associated with altered dynamic connectivity"

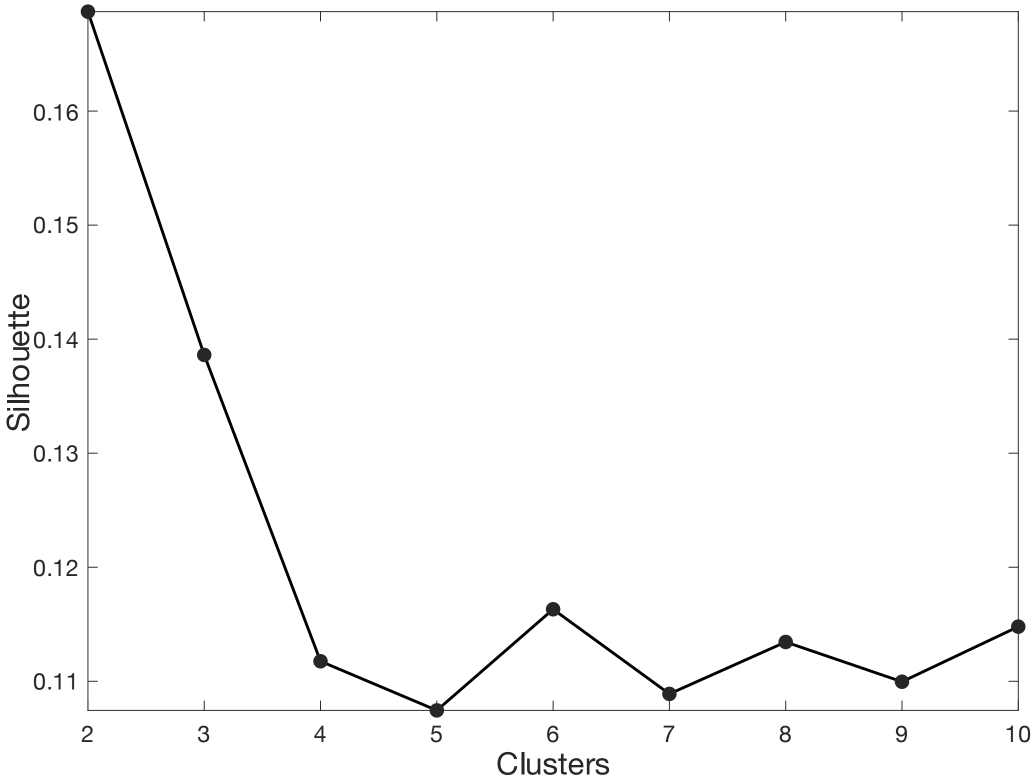


**Figure S1. Silhouette score.** The visualization of the optimal number of states determined based on the silhouette score of the cluster validity index. We here measured distance as Euclidian distance. The highest silhouette value was achieved with $k$ = 2 states.

**Table S1. Overview of Amyloid and Tau abnormalities among AD patients (N = 13)**

| Patient | Amyloid + (CSF) | Tau + (CSF) | Amyloid+ (PET) | Tau+ (PET) |
|---|---|---|---|---|
| 1 | + | + | NA | + |
| 2 | + | + | NA | + |
| 3 | + | + | NA | + |
| 4 | + | + | NA | + |
| 5 | + | + | NA | + |
| 6 | + | + | NA | + |
| 7 | + | + | NA | + |
| 8 | + | + | NA | + |
| 9 | NA | NA | + | + |
| 10 | + | + | NA | + |
| 11 | + | + | + | + |
| 12 | + | + | NA | + |
| 13 | + | + | NA | + |

The table illustrates the status of amyloid and tau biomarkers for each patient enrolled in this study. Results are reported separately for abnormalities in amyloid and tau load as assessed in cerebrospinal fluid (CSF) and in positron emission tomography (PET). A '+' indicates an abnormal value in the conducted examination. 'NA' indicates that the corresponding diagnostics were not performed. All patients tested positive for amyloid and tau with respect to at least one of the two diagnostic modalities.

**Table S2A. Results of the neuropsychological assessment in the study population.**

| Subtest | Number of impaired patients | AD patients | Healthy controls | $p_{FDR}$ |
|---|---|---|---|---|
| Dementia Detection (DemTect) [points] (Cut-Off: < 13 points) | 11 | 8.92 ± 3.43 (N = 13) | 13.92 ± 1.66 (N = 13) | **< 0.001*** |
| Cologne Neuro-psychological Screening for Stroke Patients (KöpSS) [points] (Cut-Off: ≤ 98 points) | 11 | 81.00 ± 9.89 (N = 12) | 100.73 ± 5.68 (N = 13) | **< 0.001*** |
| Aphasia Check List Short (ACL-K) [points] (Cut-Off: < 33 points) | 4 | 33.69 ± 3.64 (N = 13) | 38.00 ± 1.35 (N = 13) | **< 0.01*** |
| Trail Making Test A [s] (Cut-Off: > 100 seconds) | 2 | 64.30 ± 33.40 (N = 12) | 36.63 ± 14.14 (N = 13) | **< 0.05*** |
| Trail Making Test B [s] (Cut-Off: > 300 seconds) | 5 | 222.00 ± 84.21 (N = 11) | 96.83 ± 58.87 (N = 13) | **< 0.001*** |
| Trail Making Test ratio [B / A] | | 3.85 ± 1.50 (N = 11) | 2.58 ± 0.95 (N = 13) | **< 0.05*** |
| Beck Depression Inventory II (BDI-II) [points] (Cut-Off: ≥13 points) | 1 | 6.27 ± 4.61 (N = 11) | 3.00 ± 3.67 (N = 13) | 0.072 |
| Montgomery Åsberg Depression Rating Scale (MADRS) [points] (Cut-Off: ≥7 points) | 3 | 4.83 ± 3.71 (N = 12) | 2.39 ± 2.76 (N = 14) | 0.072 |

The table depicts the results of the neuropsychological assessments. The results are separately displayed for the group of individuals with Alzheimer's disease and healthy controls. The number of affected patients per test is reported. Descriptive data for each group are provided as means ± standard deviations, and p-values of statistical group differences as calculated by Student's t-tests are reported for each of the subtests. All p-values were FDR-corrected for multiple comparisons.

**Table S2B. Results of the motor assessment in the study population.**

| Subtest | AD patients | Healthy controls | $p_{FDR}$ |
|---|---|---|---|
| Maximum grip strength left [kPa] | 60.39 ± 23.15 (N = 13) | 65.69 ± 21.08 (N = 12) | 0.710 |
| Maximum grip strength right [kPa] | 63.62 ± 23.63 (N = 13) | 68.50 ± 21.02 (N = 12) | 0.710 |
| Maximum grip strength ratio [left / right] | 0.95 ± 0.10 (N = 13) | 0.96 ± 0.14 (N = 12) | 0.868 |
| Maximum finger tapping frequency left [Hz] | 5.21 ± 1.32 (N = 12) | 5.83 ± 0.81 (N = 12) | 0.360 |
| Maximum finger tapping frequency right [Hz] | 5.53 ± 1.93 (N = 12) | 6.08 ± 1.01 (N = 12) | 0.587 |
| Maximum finger tapping frequency ratio [left / right] | 1.02 ± 0.98 (N = 12) | 0.98 ± 0.18 (N = 12) | 0.753 |
| Purdue pegboard left [# pegs] | 10.10 ± 1.81 (N = 13) | 11.33 ± 2.43 (N = 13) | 0.360 |
| Purdue pegboard right [# pegs] | 10.75 ± 2.14 (N = 13) | 12.64 ± 2.06 (N = 13) | 0.198 |
| Purdue pegboard ratio [left / right] | 0.95 ± 0.09 (N = 13) | 0.89 ± 0.11 (N = 13) | 0.360 |
| Jebsen-Taylor Test left [s] | 36.61 ± 9.06 (N = 13) | 32.93 ± 9.52 (N = 13) | 0.554 |
| Jebsen-Taylor Test right [s] | 39.93 ± 14.38 (N = 13) | 30.41 ± 4.74 (N = 13) | 0.198 |
| Jebsen-Taylor Test ratio [left / right] | 0.96 ± 0.18 (N = 13) | 1.07 ± 0.16 (N = 13) | 0.360 |
| Action Research Arm Test (ARAT) left | 56.92 ± 0.28 (N = 13) | 57.00 ± 00.00 (N = 13) | |
| Action Research Arm Test (ARAT) right | 56.92 ± 0.28 (N = 13) | 57.00 ± 00.00 (N = 13) | |
| Action Research Arm Test (ARAT) [left / right] | 1.00 ± 00.00 (N = 13) | 1.00 ± 0.00 (N = 13) | |

The table depicts the results for each subtest comprising the motor assessment. The results are displayed for the entire study population, divided into the group of individuals with Alzheimer's disease and healthy controls. Descriptive data for each group are provided as means ± standard deviation (SD), and p-values of statistical group differences as calculated by Student's t-tests are reported for each of the subtests. All p-values were FDR-corrected for multiple comparisons.
Since healthy controls performed at ceiling and without any variance for the Action Research Arm Test (ARAT), no group comparison could be performed for the parameters of the ARAT.
